# DEVELOPMENT AND VALIDATION OF A PREDICTION RULE FOR BENEFIT AND HARM OF ORAL ANTICOAGULATION IN NON-VALVULAR ATRIAL FIBRILLATION

**DOI:** 10.1101/2024.06.20.24309269

**Authors:** Sergio Raposeiras-Roubin, Tze-Fan Chao, Emad Abu-Assi, Yi-Hsin Chan, Inmaculada González Bermúdez, Jo-Nan Liao, Ling Kuo, Rocío González Ferreiro, Andrés Íñiguez-Romo

## Abstract

**Background:** Oral anticoagulation therapy (OAC) remains the gold standard for stroke prevention in patients with atrial fibrillation (AF). In real life, there are patients who do not receive OAC due to high bleeding risk. In those patients, left atrial appendage closure (LAAC) has emerged as a potential alternative for stroke prevention. With this study, we aimed to develop a clinical decision tool to identify patients expected to derive harm vs benefit from OAC therapy.

**Methods:** Among 14,915 AF patients with CHA2DS2-VASc ≥ 1 (2 for women) from CardioCHUVI-AF registry (78% with OAC), a prediction rule was derived using a linear regression model to predict the stroke-bleeding balance. This rule was externally validated in the Taiwan AF registry, with 26,595 patients (70.5% with non OAC therapy).

**Results:** A simplified risk score was created using 7 clinical variables. The low-score group (≤ −8 points) was associated with higher rates of bleeding than stroke (7.25 vs 1.11 and 3.27 vs 2.58 per 100 patients/year in derivation and validation cohorts, respectively; p<0.001). In those patients, OAC was harmful. In contrast, high-score group (≥ +6 points) was associated with higher stroke risk than bleeding risk (2.32 vs 1.71 and 4.19 vs 1.64 per 100 patients/year in derivation and validation cohorts, respectively; p<0.001), with great benefit of OAC therapy.

**Conclusions:** A prediction rule balancing stroke and bleeding risks correctly identify patients with harm vs benefit from OAC therapy. This rule requires further prospective evaluation to assess potential effects on patient care to select candidates for LAAC.

**CLINICAL PERSPECTIVE:** *What is new?:*

- We developed and externally validated a simple user-friendly clinical tool -OAC score- to balance both embolic and bleeding risks in atrial fibrillation patients with CHA2DS2-VASC ≥1 (excluding female sex).
- According to OAC score, we can identify atrial fibrillation patients expected to derive benefit vs harm from anticoagulation therapy.
- Patients with a low OAC score have much higher rates of bleeding than stroke, which is exaggerated with anticoagulation. The opposite occurs in patients with a high score.

*What are the clinical implications?:*

- A standardised stratification of both stroke and bleeding risk in atrial fibrillation patients at high embolic risk can be performed using an user-friendly, comprehensive tool (the OAC score).
- In patients with low risk OAC score, despite high risk CHA2DS2-VASC, anticoagulation is not beneficial and left atrial appendage closure could be an alternative.
- Further studies are needed to assess the impact of oral anticoagulation and left atrial appendage closure in patients with atrial fibrillation according to the OAC score risk groups.

## INTRODUCTION

The CHA_2_DS_2_-VASc score has a great clinical impact to select patients with atrial fibrillation (AF) at high embolic risk in order to establish the indication for oral anticoagulation (OAC)(1, 2). Use of OAC in patients with ≥ 1 non-sex CHA_2_DS_2_-VASc risk factor significantly reduces the chances of thromboembolic cardiovascular events, such as stroke, and mortality in AF patients(3). However, the clinical benefit of anticoagulant treatment is offset by the risk of bleeding. Data from the PREFER-AF(4), GARDFIELD-AF(5), and NCDR-PINNACLE(6) studies, show that 20%, 30%, and 40% of the patients with AF and a CHA_2_DS_2_-VASc score ≥2 are not treated with OAC, respectively. A perceived increased bleeding risk is the most frequent reason for withholding OAC for embolic prevention in non-valvular AF(7). Although guidelines recommend the use of bleeding risk scores −e. g. HAS-BLED−, those scores do not determine OAC prescription; they only help to identify potentially reversible bleeding risk factors in order to correct them(8).

Due to an overlap of risk factors for bleeding and stroke, patients with AF are more usually at an increased bleeding risk (9). Decision-making regarding anticoagulation can be particularly challenging in these patients, especially when both stroke and bleeding risks are high. Left atrial appendage closure (LAAC) could be a reasonable option for those patients, especially when the expected risk associated with OAC is greater than the expected benefit (10). To guide clinical decision-making for patients at high risk of stroke and AF, we designed this study focused on identifying factors that predict whether the expected increase in MB associated with OAC would outweigh the expected benefit of reduced IS for individual patients. Those factors will be used to develop a decision tool to help select AF patients without net benefit from OAC who are candidates for LAAC.

## METHODS

### Study population

A retrospective registry-based cohort study including all consecutive patients (n=16,202) with a diagnosis of AF between January 2014 and January 2018 in the health area of Vigo (Galicia, Spain) was used (*CardioCHUVI-AF* registry; ClinicalTrials.gov identifier: NCT04364516)(11). Patients were identified through administrative databases at both the hospital and ambulatory level, using the regional electronic healthcare records system. Electronic medical records were reviewed in all patients, and diagnosis of AF was confirmed by a compatible electrocardiogram. Clinical, laboratory, and therapeutic data were collected using an encoded database. Patients with missing baseline and follow-up data were excluded (n=146), together with those with valvular AF (moderate-severe mitral stenosis or mechanical prothesis valve; n=350). Patients without none non-sex CHA_2_DS_2_-VASc risk factor (n=791) were also excluded. Therefore, the final cohort comprised 14,915 AF patients with CHA_2_DS_2_-VASc ≥ 1 for men and ≥ 2 for women. The study was conducted in accordance with the principles of the Declaration of Helsinki and was approved by the local ethics committee (*Autonomous Committee of Research Ethics of Galicia*, code HAC-ACO-2018-01, registry 2018/258).

### Study Goals

The goal of this study was to identify clinical variables that determine whether a patient’s bleeding risk is lower or higher than the embolic risk, in order to distinguish patients who can derive the greatest benefit from those who can experience the most harm from OAC therapy. Considering individual patient characteristics and their independent associations with Ischemic Stroke (IS) and Major Bleeding (MB), this study sought to stratify outcomes based on a single multivariable risk score. This entailed (1) identifying factors associated with IS and MB, (2) choosing those that selectively predicted either IS or MB to generate a simplified risk score, and (3) assessing the OAC therapy results observed in the registry, stratified by the new risk score. An ideal score would identify patients with simultaneous high stroke risk (and corresponding high benefit with OAC therapy) and low bleeding risk (and corresponding low risk of harm with OAC therapy), and vice versa. In addition, the ability of the score to balance both IS and MB risks within an external sample was assessed, using the Taiwan AF registry with 26,595 patients with AF and CHA_2_DS_2_-VASc score ≥ 1 for men or ≥ 2 for women.

### Embolic and Bleeding End Points

The primary embolic end point was IS (as defined by the Standardized Data Collection for CV Trials Initiative and the US Food and Drug Administration)(12), and the primary bleeding end point was MB (as defined by the International Society on Thrombosis and Haemostasis)(13). During the follow-up, two data managers independently reviewed the clinical history of each patient to identify possible events. All suspected events were independently reviewed by 2 specialists in the clinically relevant areas of cardiology and internal medicine. Both data managers and specialists underwent a study-specific training on event definitions and the adjudication process. Finally, an independent clinical events committee revised and adjudicated all events.

### Statistical Analysis

#### Development of Stroke and Bleeding Risk Models

Clinical characteristics were compared between patients experiencing events during follow-up and those without events, using Fisher exact or t tests as appropriate. Cox regression was used to develop 2 separate models: the first to predict IS and the second to predict MB. Data were censored at the time of IS or MB, or at the time of death, change in OAC therapy status, or last known contact, whichever was earliest. A total of 40 candidate variables potentially associated with IS or MBs based on a comprehensive literature review and clinical plausibility were identified (Table 1). Candidate variables that differed in bivariable comparisons at a significance level ≤ 0.150 were incorporated. Stepwise selection was then performed, using the 0.050 significance level. To identify possible heterogeneous treatment effects, simple Cox regression models were developed for each outcome including treatment group, variable of interest, and their interaction term. Interactions terms significant at a P value ≤ 0.100 were entered into the stepwise selection process with other candidate variables. Proportionality was evaluated for all variables in the models. Model discrimination was assessed using Harrell’s c-statistic, and calibration with the Gronnesby and Borgan goodness-of-fit test.

**Table 1.**
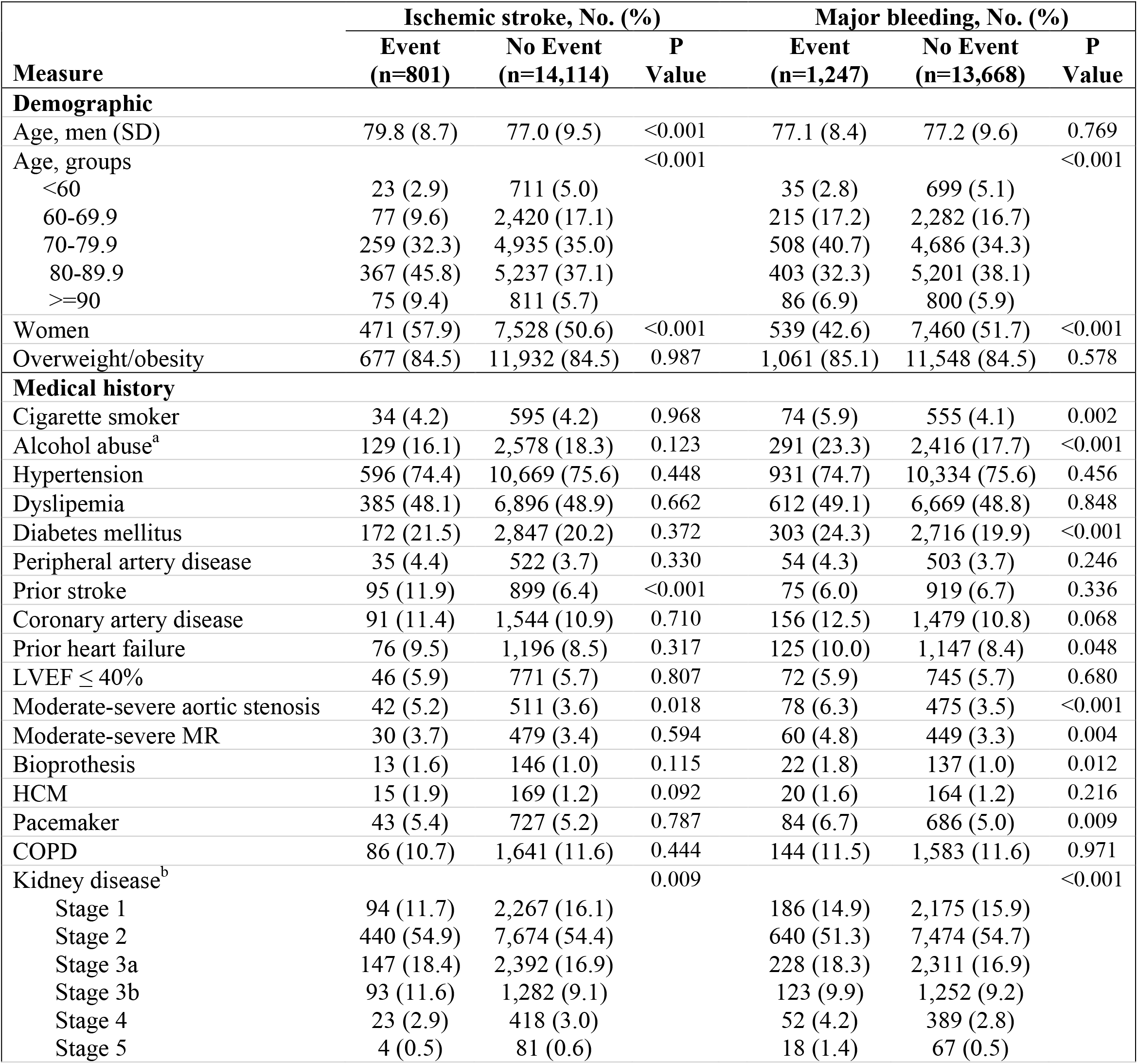
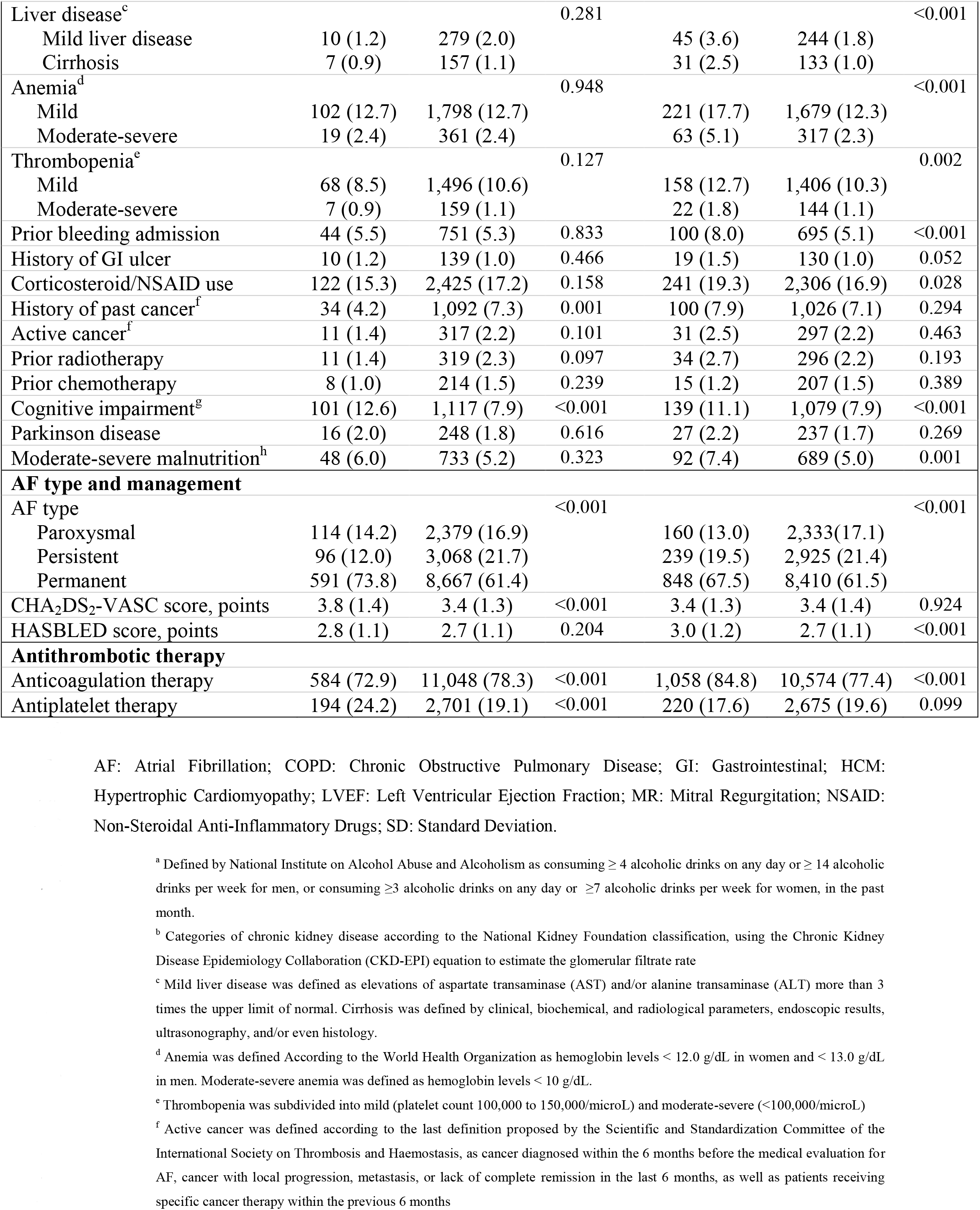
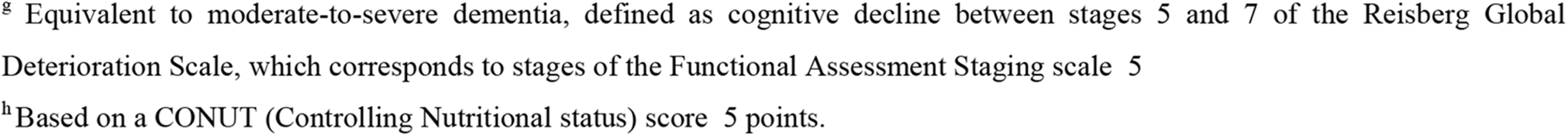
Baseline characteristics of patients with vs without ischemic stroke or major bleeding during the follow-up in the derivation cohort (n = 14,915)

### Development of a Simplified Clinical Prediction Score. For each patient, the predicted risks

(cumulative incidence) of IS and MB were estimated, assuming OAC therapy and separately assuming non-OAC therapy. The difference between these 2 predicted values represented the predicted balance between IS and MB. A linear regression model was created, using the predicted balance between IS and MB −as the outcome− and all predictors that were selected in the stroke and bleeding models. Baseline variables that statistically accounted for more than 1% of the observed variation in estimated balance between IS and MB were included in a simplified clinical prediction score, called OAC score. All variables were assigned an integer score of 1 or 2 (or −1 to −2) based on β coefficients. The range of potential scores was between −22 and 10. Clinically relevant score groupings were created, defining patients more likely to be harmed from OAC (low score group) vs those more likely to benefit (moderate-high score groups). Low score was defined as percentile 10 of patients with negative OAC score values (i.e. a score ≤ - 8), and high score as percentile 90 of patients with positive OAC score values (i.e. a score ≥ 6). Kaplan-Meier event rates were compared by score groups, and absolute risk differences in IS and MB rates across score groups were assessed. Statistical analyses were conducted using *STATA* software, version 15 (Stata Corp, College Station, Texas, USA).

## RESULTS

### Derivation cohort

A total of 14,915 patients with AF and CHA_2_DS_2_-VASc ≥1 for men or ≥2 for women were included in this analysis (derivation cohort). Of these, 11,632 patients (78.0%) receiving OAC (86.8% VKA, 13.2% DOAC). During a mean follow up of 4.0 years (IQR 2.4-5.7 years), 801 patients developed IS (1.3 per 100 patients/year) and 1,247 patients developed MB (2.1 per 100 patients/year). Patients who had an IS during follow-up were older, more likely women, had higher rates of prior stroke, aortic stenosis, hypertrophic cardiomyopathy (HCM), chronic kidney disease (CKD), history of cancer and dementia, and were less likely to have been treated with OAC compared with patients without an IS (Table 1). Patients with a MB were also older, more likely men, had a higher prevalence of smoking, enolism, and diabetes, had a higher prevalence of cardiovascular disease (including prior heart failure and valvular heart disease), higher comorbidity (including dementia, malnutrition, liver disease, CKD, anemia, trombopenia and prior bleeding admission), and were more likely to have treated with OAC, corticosteroids and NSAIDs compared with patients without a MB (Table 1).

### Risk Prediction Models

In multivariable Cox regression (Table 2), significant predictors of both IS and MB included age, aortic stenosis, dementia, and permanent AF. Variables that predicted only the risk of IS included prior stroke and HCM. Male sex, smoking, diabetes mellitus, mitral regurgitation, CKD, liver disease, anemia, prior bleeding admission, prior gastrointestinal ulcer, malnutrition, and corticosteroid/NSAID use were significant independent predictors of MB, but not of IS. Both models had moderate discrimination (c statistic 0.687 and 0.671 for stroke and bleeding models, respectively), and were well calibrated (goodness-of-fit P = 0.116 and 0.549 for stroke and bleeding models, respectively).

**Table 2.**
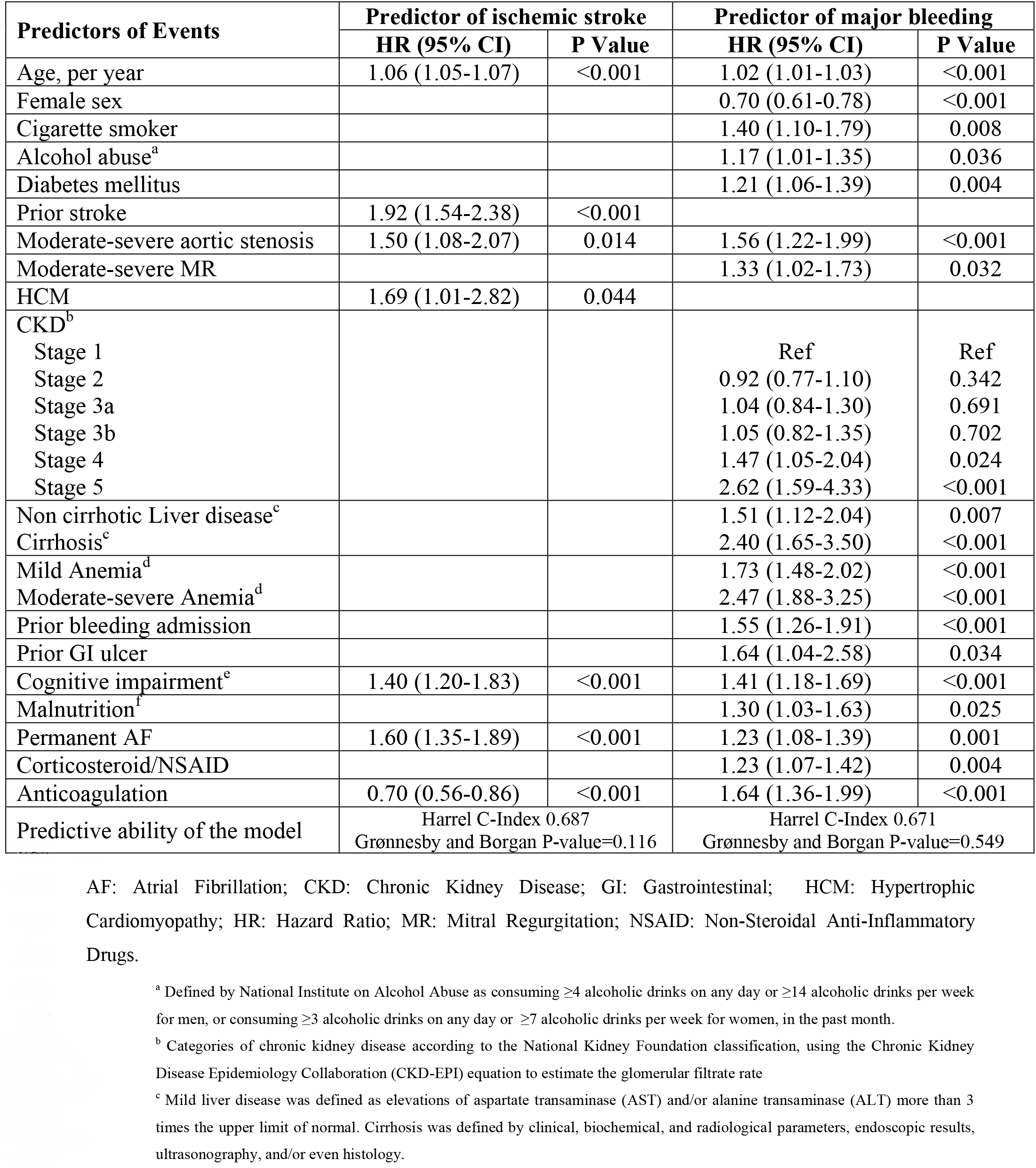
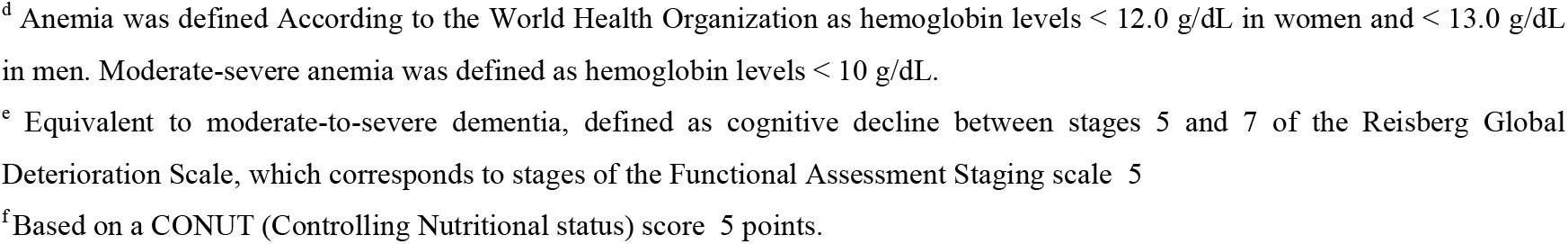
Ischemic stroke prediction model and major bleeding prediction model.

### Clinical Prediction Score

A simplified risk score −called OAC score− was created using 7 clinical variables (age, sex, prior stroke, prior bleeding, anemia, renal function, and liver function) that were independently associated with more than 1% of the variation in the balance between IS and MB in a linear regression model (Supplementary Table 1). The score, ranging from −22 to 10, assigned points as follows: for patients younger than 60 years, 0 points; for age 60 to 69,+1; for age 70 to 79,+2; for age 80 to 89,+3; for patients 90 years or older,+4; for female sex, +3; for prior stroke, +4; for CKD stage 5, −11; for non-cirrhotic liver disease (ALT or AST > 3 USL), - 6 points; for cirrhotic liver disease, −10 points; for hemoglobin 10 to 11.9 g/dL, −5 points; for hemoglobin lower than 10 g/dL, −9 points; for prior bleeding admission, −4 points. Patients were classified in 3 risk groups according to OAC score: low (≤ - 8 points; n=242, 1.6%), moderate (from −8 to +6 points; n=11,264, 75.5%), and high score (≥ + 6 points; n=3,409, 22.9%). The low-score group was associated with the highest rates of MB and the most negative balance between IS and MB, whereas the opposite occurred in the high-score group (Figure 1). Kaplan Meier curves for MB and IS according the 3 risk groups are showed in Figure 2A and 2B. The impact of OAC therapy in stroke-bleeding balance is shown in Figure 3A.

**Figure 1.**
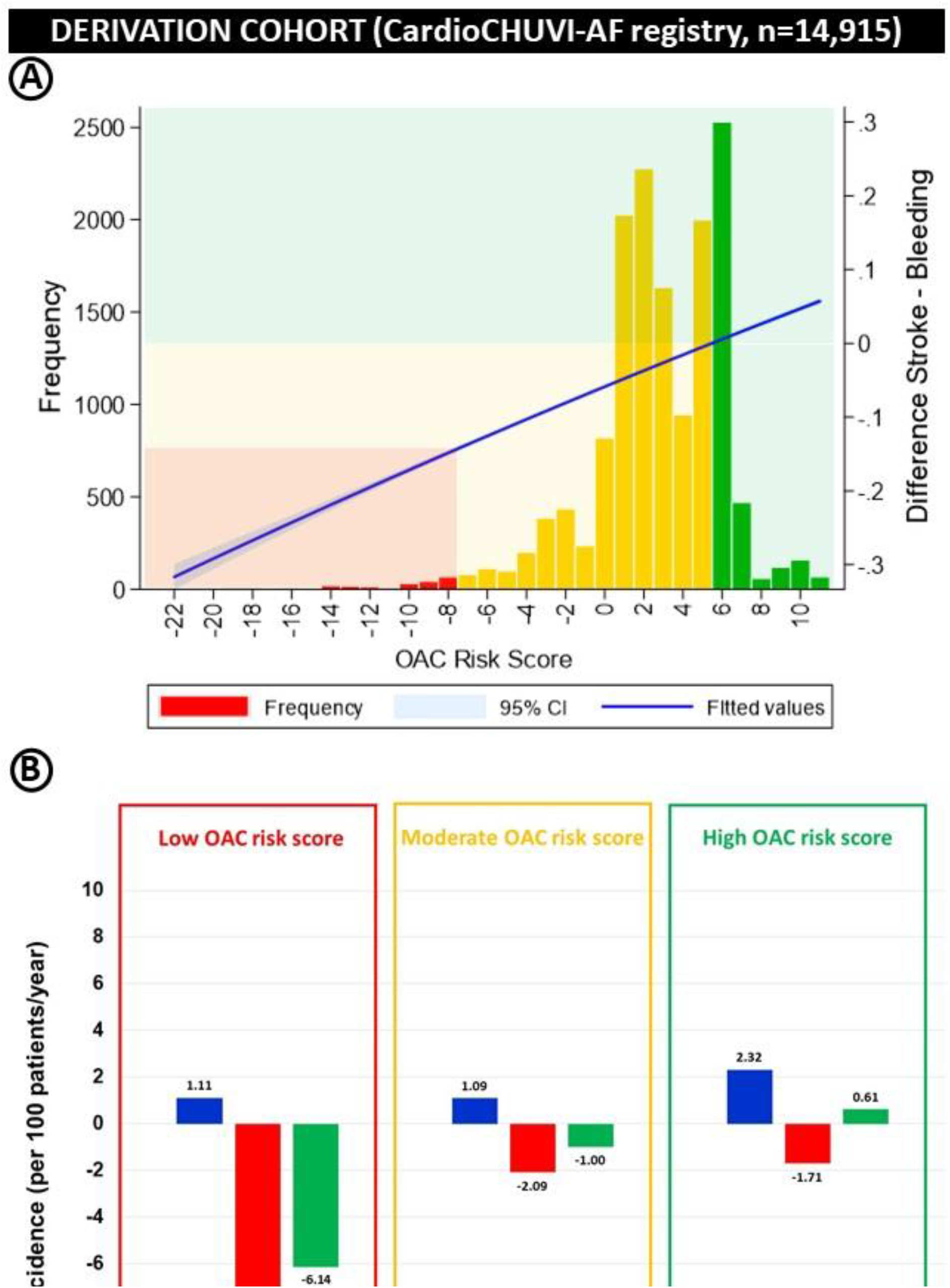
Distribution of OAC score among patients of derivation cohort (n = 14,915) with annual incidence of events according to risk score groups.

**Figure 2.**
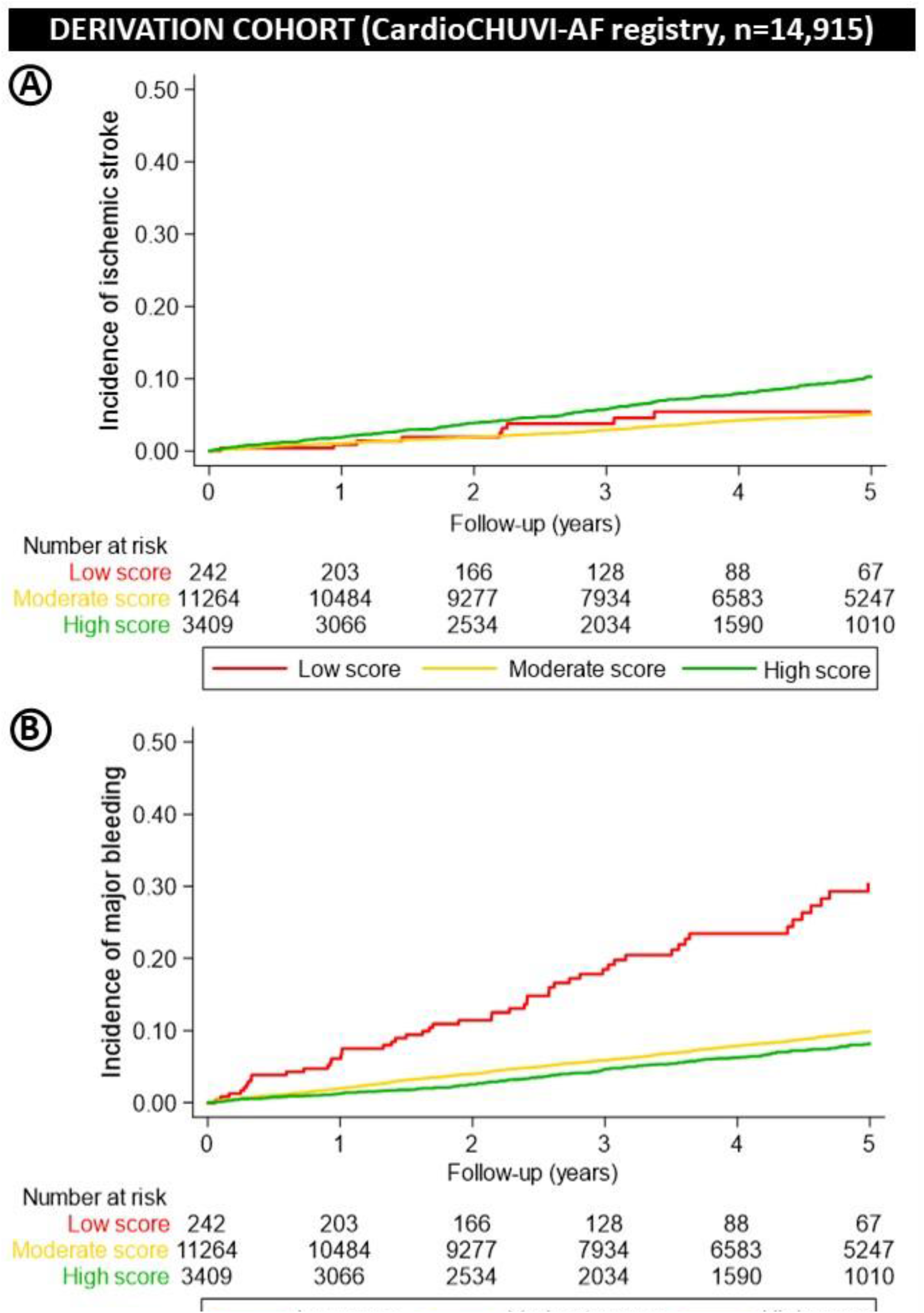
Observed rates of embolic (A) and bleeding (B) outcomes during the follow-up by OAC score groups in derivation cohort.

**Figure 3.**
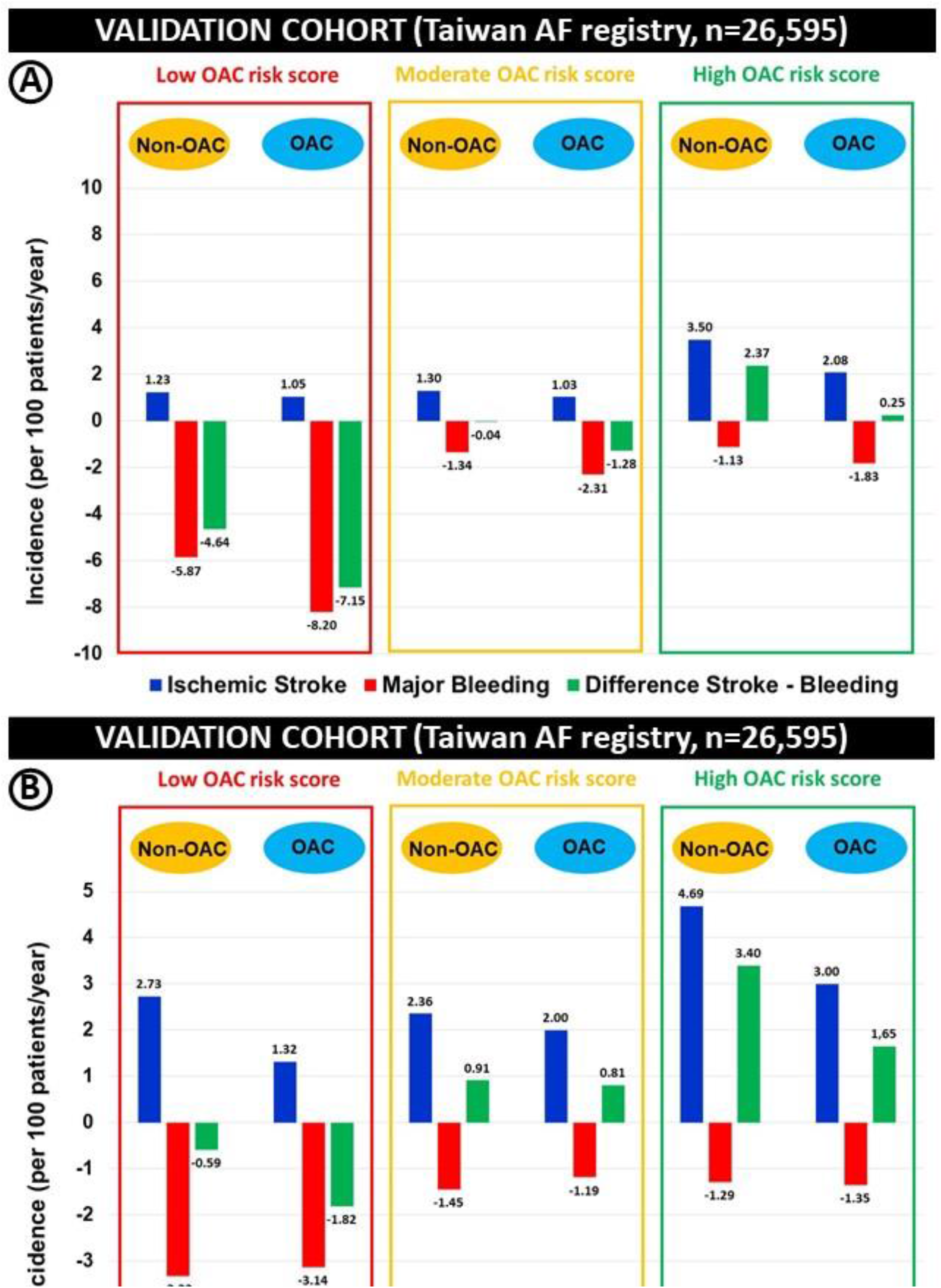
Balance between embolic and bleeding outcomes in anticoagulated and non-anticoagulated patients according to OAC score groups in both derivation and validation cohorts.

### External validation of OAC score

OAC score was validated in a Nationwide Cohort Study from Taiwan, using the National Health Insurance Research Database in Taiwan(14). Among the patients of Taiwan AF registry with CHA2DS2-VASc ≥ 1 (2 for women), data of OAC score were available in 26,595 patients. From them, only 29.5% were treated with OAC therapy. OAC risk score was independently associated with the balance of embolic-hemorrhagic risk (Figure 4). According to the OAC score, 19.7%, 74.0%, and 6.3% of patients are classified as low, moderate, and high score. A high OAC score is associated with more IS whereas a low OAC risk score is associated with more MB (Figures 5A and 5B). Only patients with a low OAC risk score had a negative embolic-hemorrhagic balance, that was exaggerated in anticoagulated patients (Figure 3B).

**Figure 4.**
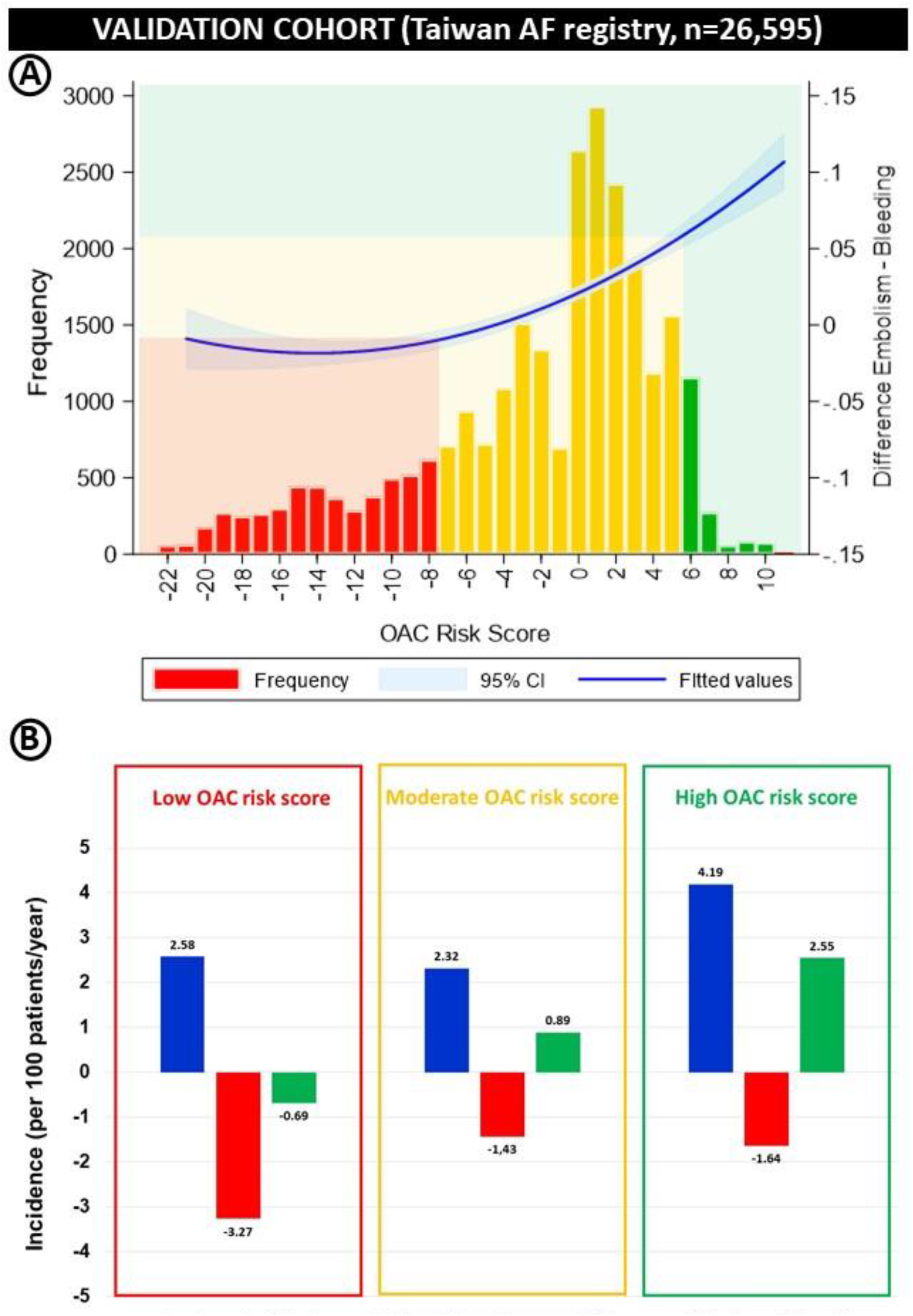
Distribution of OAC score among patients of validation cohort (n = 26,595) with annual incidence of events according to risk score groups.

**Figure 5.**
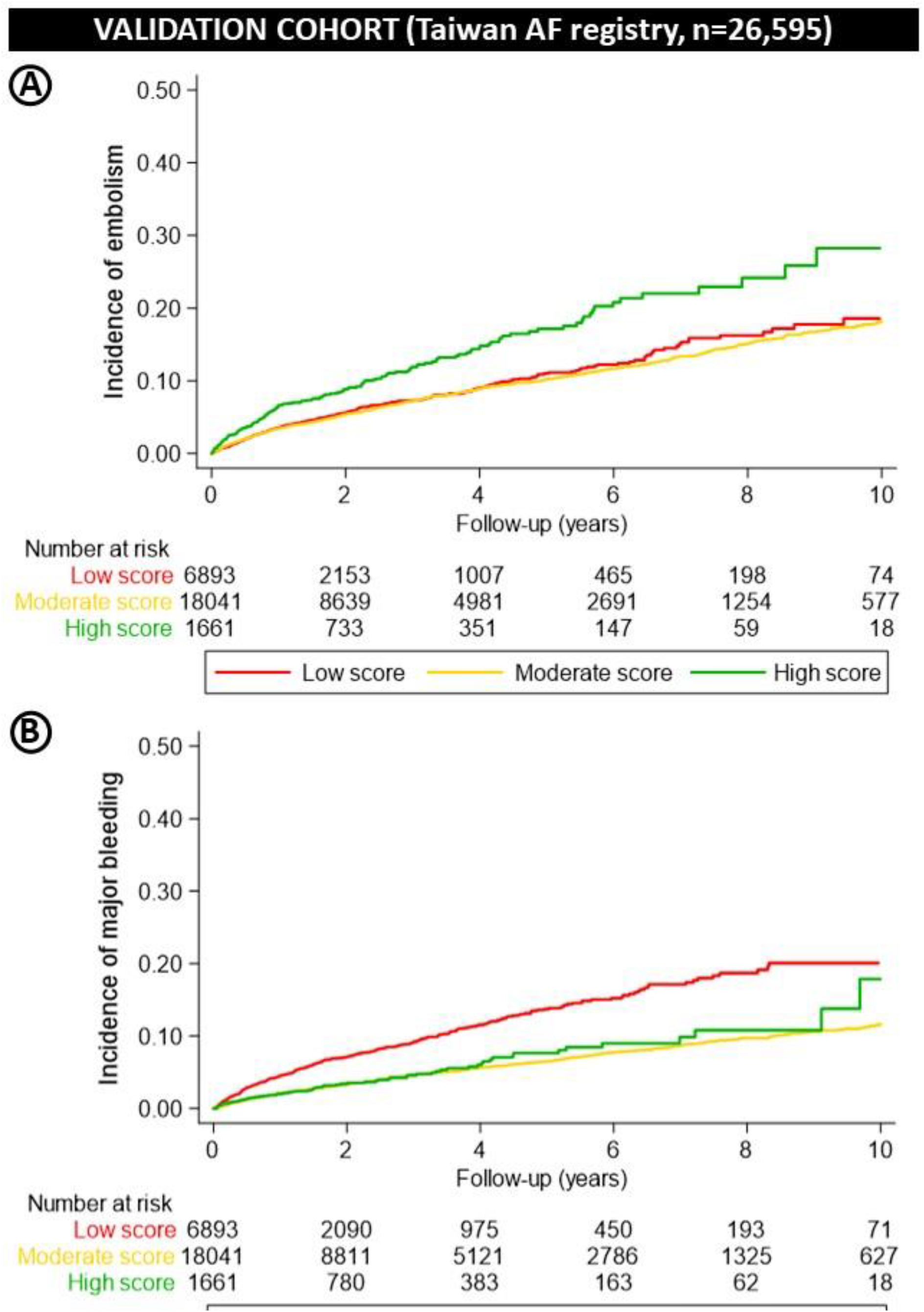
Observed rates of embolic (A) and bleeding (B) outcomes during the follow-up by OAC score groups in validation cohort.

## DISCUSSION

This study developed and validated a clinical prediction score based on embolic and bleeding risk factors to help physicians to identify AF patients with greater expected harm vs greater expected benefit from OAC therapy. Despite prior evidence suggesting that embolic and bleeding risk are strongly correlated, our results suggest that it may be possible to identify individual patients with discordant embolic risks and bleeding risks.

In patients with a low OAC score (i.e, ≤ −8 points), the risk of bleeding events was clearly higher than the risk of embolic events in both the derivation cohort (78.0% with OAC therapy) and the validation cohort (70.5% without OAC therapy). This finding was exaggerated in anticoagulated patients. This is very interesting, because we can identify a group of patients in who the benefit of OAC therapy against embolism is compromised by the increased bleeding risk. In those patients, an alternative strategy should be sought, like LAAC. A meta-analysis of 2 randomized clinical trials (PROTECT AF and PREVAIL) comparing LAAC vs anticoagulation with vitamin K antagonists (VKA) in 2,406 patients at high risk for stroke and increased risk of bleeding, showed a significant reduction of almost 50% in nonprocedural bleeding (15). In turn, LAAC was associated with similar rates of nonprocedural stroke, and resulted in improved rates of hemorrhagic stroke and cardiovascular/unexplained death. Recently, data from PRAGUE-17 showed that among patients at high risk for stroke and increased risk of bleeding, LAAC vs direct oral anticoagulants (DOACs) did not increase the rate of stroke (HR 1.00, 95%CI 0.40-2.51; p=0.99), whereas showed a trend to decrease the rate of nonprocedural bleeding (HR 0.53, 95%CI 0.26-1.06; p=0.07) (10). Current guidelines state that LAAC may be considered in AF patients with contra-indications for long-term OAC therapy. However, with the results of PROTECT-AF, PREVAIL and PRAGUE-17, it seems reasonable to take a step forward and recommend it in those patients in whom the bleeding risk clearly exceeds the embolic risk. In this sense, the OAC score can be very useful for clinicians, helping to the incorporation of transcatheter LAAC into a stroke prevention strategy for patients with an expected excess bleeding rate. OAC score considers the balance between two key elements: the long-term risks of thromboembolic events and the longer-term risk of bleeding. However, for derivation to LAAC, another key element should be considered: the short-term procedure risk of LAAC(16). With that in mind, the risk benefit of an invasive procedure such as LAAC must be balanced against the longer-term issues of continued exposure to anticoagulation and bleeding(17).

Other therapeutic option to prevent stroke in patients with high bleeding risk could be Factor XIa (FXIa) inhibitors. Whether FXIa inhibitors can play a role in this clinical scenario is currently a pending challenge. FXIa inhibition represents a promising strategy to reduce stroke risk while preserving a patient’s ability to clot when bleeding(18). The recent PACIFIC-AF trial showed that asundexian −a FXIa inhibitor− resulted in lower rates of bleeding compared with apixaban in patients with AF, with similar embolic protection(19). These data support the potential role for FXIa inhibition in patients with AF and make this strategy a promising avenue pending the results of future studies.

On the other hand, a high OAC score identifies patients with a very high embolic risk that markedly exceeds the expected risk of bleeding events, both in anticoagulated and non-anticoagulated patients −an aspect that is fulfilled in both derivation and validation cohorts−. In those patients there is a clear benefit of anticoagulation, and even indefinite OAC with LAAC could be useful, giving in consideration that the left atrial appendage is responsible for >90% of embolic strokes(20). Although a targeted approach to exclude this chamber combined with OAC therapy can be beneficial in those patients at very high embolic risk(21), careful consideration of LAAC warrants an individualized discussion between physician and patient.

### Limitations

A number of limitations should be considered in interpreting these findings. As in any observational cohort study, our findings may reflect some degree of residual confounding. First, as it is not a randomized clinical trial, there is a clear selection bias when deciding to prescribe OAC. Therefore, these data could only be used to evaluate whether the score stratified patients according to the balance between embolic and bleeding risk, and not actual benefit or harm with OAC therapy. The usefulness of the OAC score to support clinical decision on the prescription of OAC therapy or LAAC should be regarded as hypothesis generating at this stage and dedicated randomized trials should be carried out to evaluate the validity of this potential application of the OAC score. Second, variables in the predictive score included patient characteristics that have demonstrated an association with either embolic or bleeding events in prior studies. However, common predictive factors for both embolic and bleeding risks were not included in the predictive score because these factors did not help identify discordant bleeding and embolic risks. The incorporation of more variables into the individual bleeding and embolic models may have improved discrimination, at the expense of parsimony. In addition, the estimation of risks based on the use of the separate embolic and bleeding model coefficients rather than use of the simplified score could improve the ability to predict such events and provide the opportunity for clinicians to identify patients with concordantly high embolic and bleeding risks, in addition to those with discordant risks. However, we have searched for a simple and user-friendly score that has clinical implications and widespread use. Third, although there remains a sizable proportion of patients, the patients used to derive the OAC score make up a group of patients −most of them anticoagulated− that may not be representative of those seen in clinical practice. However, validation was performed in a different population of AF patients −most of them non-anticoagulated− with concordant results. Despite our optimism, use of this prediction score should be cautious until further validation is performed in clinical trials, and optimal clinical and procedural care to reduce overall bleeding and ischemic risks should be practiced independent of a patient’s score.

In conclusion, we have developed and tested the OAC score, a user-friendly tool to balance bleeding and embolic risks in AF patients. OAC score is feasible and effective with potentially important implications on the selection of patients for LAAC.

## Data Availability

Data supporting this study are available from CardioChuvi registry. Access to the data is subject to approval and a data sharing agreement due to local policies

## Contributors

- SRR, TFC, EAA, and AIR conceived and designed the study.
- IGF, RGF, YHC, JNL, and LK a did the analysis.
- SRR, EAA and TFC drafted the manuscript.
- All authors revised and approved the final version of the manuscript.
- SRR and EAA had full access to all of the data in the study and take responsibility for the integrity of the data and the accuracy of the data analysis.

## Founding source

- The electronic registry was funded in a unconditioned way by Daiichi Sankyo, Pfyzer-BMS, Bayer and Boehringer Ingelheim.

## Acknowledgment

None.

## Data sharing

- Due to different data-sharing policies of the two datasets included in this study, not all of which provided free access to data, data included in this study will not be made available. Requests for the data from each included dataset should be made to the corresponding author of each single registry.

## ABBREVIATIONS

AF: Atrial Fibrillation
CKD: Chronic Kidney Disease
HCM: Hypertrophic Cardiomyopathy
IS: Ischemic Stroke
LAAC: Left Atrial Appendage Closure
MB: Major Bleeding
OAC: Oral Anticoagulation

